# Patient acceptability of ctDNA testing in endometrial cancer follow-up

**DOI:** 10.1101/2020.07.15.20154195

**Authors:** A Relton, A Collins, DS Guttery, D Gorsia, HJ McDermott, EL Moss

## Abstract

**Objective:** Circulating tumour DNA (ctDNA) is emerging as a potential option to detect disease recurrence in many cancer types however, ensuring patient acceptability of changing clinical practice and the introduction of new technology is paramount. This study aimed to explore women’s opinions on the acceptability of ctDNA to monitor for endometrial cancer (EC) recurrence.

**Methods:** Women enrolled on a non-intervention cohort study determining the ability of ctDNA to detect recurrent endometrial cancer were invited to participate in a semi-structured interview. Data was analysed using Template Analysis.

**Results:** Eighteen women were interviewed. Participants represented a mix of cases, including early stage high-risk EC, metastatic disease at diagnosis and EC recurrence, to ensure a wide range of participant experiences were captured. A ctDNA blood test was viewed by participants as more physically and psychologically acceptable than clinical examination to monitor for EC recurrence. In particular, participants expressed overwhelming preference for a blood test rather than pelvic examination. Although participants acknowledged that an abnormal ctDNA result could cause anxiety, they expressed a preference to be informed of their results, even if a recurrence was too small to detect radiologically. Explanations for these opinions were a desire for certainty whether their cancer would recur or not, and knowledge would help them be more aware of symptoms that should be reported to their clinician.

**Conclusions:** ctDNA monitoring to identify EC recurrence appears to be acceptable to patients, and for many, may be preferable to clinical examination.

## INTRODUCTION

The identification of circulating tumour DNA (ctDNA) is hailed as introducing a paradigm shift in cancer management, enabling a more personalised approach to treatment. One of the reported advantages of ctDNA is its high sensitivity and specificity in identifying patients who will experience disease relapse, often many months, or even years, before clinical signs/symptoms or detection on imaging [1, 2]. Monitoring ctDNA levels to identify cancer recurrence before it is clinically or radiologically detectable has the potential to dramatically impact patient management by being used as the basis of remote monitoring schemes, as compared to the current standard management for most tumour sites of clinician-led hospital follow-up (HFU). However, the issue of patient management and expectations with raised, or rising, ctDNA levels but no detectable site of recurrence does pose significant challenges, since earlier detection may not result in earlier oncological intervention or ultimately impact patients’ overall survival. Despite the huge international effort focusing on the scientific and molecular aspects of ctDNA detection very little work has been undertaken with the target patient groups to determine the acceptability of such developments and the potential on changing management.

Endometrial cancer (EC) is the second most common cancer in women in the USA with over 60,000 new cases diagnosed each year [3]. It has a high overall survival, 69.1-76.5% 5-year failure-free survival for high-risk disease [4], and for low-risk disease the 10-year survival rate is over 94% [5]. As a result there is a high prevalence of EC survivors in the community and this is projected to increase from 757,190 in 2016 to 942,670 over the next decade^3^, The value of HFU for early stage EC is been questioned given the low recurrence rate and that the majority of recurrences are associated with symptoms [6]. This has led to the introduction of reduced schedule or alternative models of follow-up [7], such as telephone (TFU) or patient-initiated follow-up (PIFU), which appear to be well tolerated [8, 9] and cost savings for both the patient and the healthcare economy [10]. Concerns however, have been raised over the psychological impact on patients, with fear of recurrence reported to be higher with PIFU than with HFU [11].

ctDNA is reported to have 100% sensitivity in detecting EC recurrence and a lead time of 10 months compared to CT scan and up to 18 months over patient symptoms [12]. The high accuracy of ctDNA in identifying recurrence opens up the possibility of developing remote monitoring for EC by combining ctDNA monitoring with reduced schedule follow-up, such as a PIFU scheme. This could enable the identification of women at high-risk of recurrence for further investigation and close monitoring, whilst allowing ctDNA negative women to continue on PIFU.

Before trials determining the utility of remote monitoring with ctDNA in EC can by undertaken, patient acceptability needs to be established. The aim of this study was to explore the views of study participants on the potential use of ctDNA in EC follow up in the future.

## METHODS

A prospective cohort study aimed at determining the sensitivity of ctDNA to monitor EC activity (ECctDNA study) opened for recruitment in December 2017 at the University Hospitals of Leicester. Women attending follow-up appointments for high-risk/advanced/recurrent EC had a blood sample taken for ctDNA analysis at each clinic visit. The patient and their medical team were blinded as to the ctDNA result, therefore the result did not influence the patients’ management. A qualitative study using one-to-one semi-structured interviews to investigate patient acceptability was included in the study design. Ethical approval was granted for this study by the Wales Research Ethics Committee 7 (17/WA/0342).

### Participants

Purposive sampling of the ECctDNA study population was performed in order to ensure a diverse interview study population with regards to age, ethnicity, stage at diagnosis and treatment experiences. Women attending for a routine follow-up appointment were given a printed invitation letter containing full information about the study and were verbally invited to attend for an interview. Interviews continued until data saturation was reached.

### Procedure

Data collection took place between January and June 2019. Interviews were conducted in English by the same female researcher (AR), trained in interview techniques, and independent to the ECctDNA study team. All women signed a consent form and agreed to audio recording of the interview. A topic guide (Supplementary data) was developed prior to the data collection through discussions with the ECctDNA study team, with open-ended questions and prompts to encourage discussion. The guide allowed the researcher to explore participants’ experiences with the ECctDNA blood test as an alternative to clinical examination for the detection recurrent disease. Interviews ranged from 20 minutes to 45 minutes in length and field notes were taken. The interviews were transcribed verbatim by the researcher or by an external agency. The participants did not review the transcripts or provide further comments on the results.

### Analysis

The data were analysed using Template Analysis [13]. Initially, this was deductive, whereby the interview topics formed a template and guided the analysis, but as the analysis progressed, this became more inductive in nature, as new information was added to the template [14]. Throughout development, the template was refined to give an accurate representation of themes present in the data. The transcripts were coded by the researcher and a subset were independently coded by the senior authors. Themes and codes were developed and added into a final version of the template, with codes presented and ordered to represent the different themes, both broad and specific. Evidence of the template analysis process was recorded, by maintaining copies of each stage of analysis, from the complete transcripts to each version of the developing template and final interpretation [15].

## RESULTS

In total, 18 women attended for an interview, 35% of the active ECctDNA study population. Three women declined to attend for an interview. The median age was 67.5 years and two (11.1%) of the women were of non-white ethnicity (Table 1). There was a mix of clinical cases including early stage high-risk cases, advanced disease at diagnosis and EC recurrence, to ensure a wide range of experiences. In three interviews a friend or family member was also present. There were no repeat interviews.

**Table 1.**
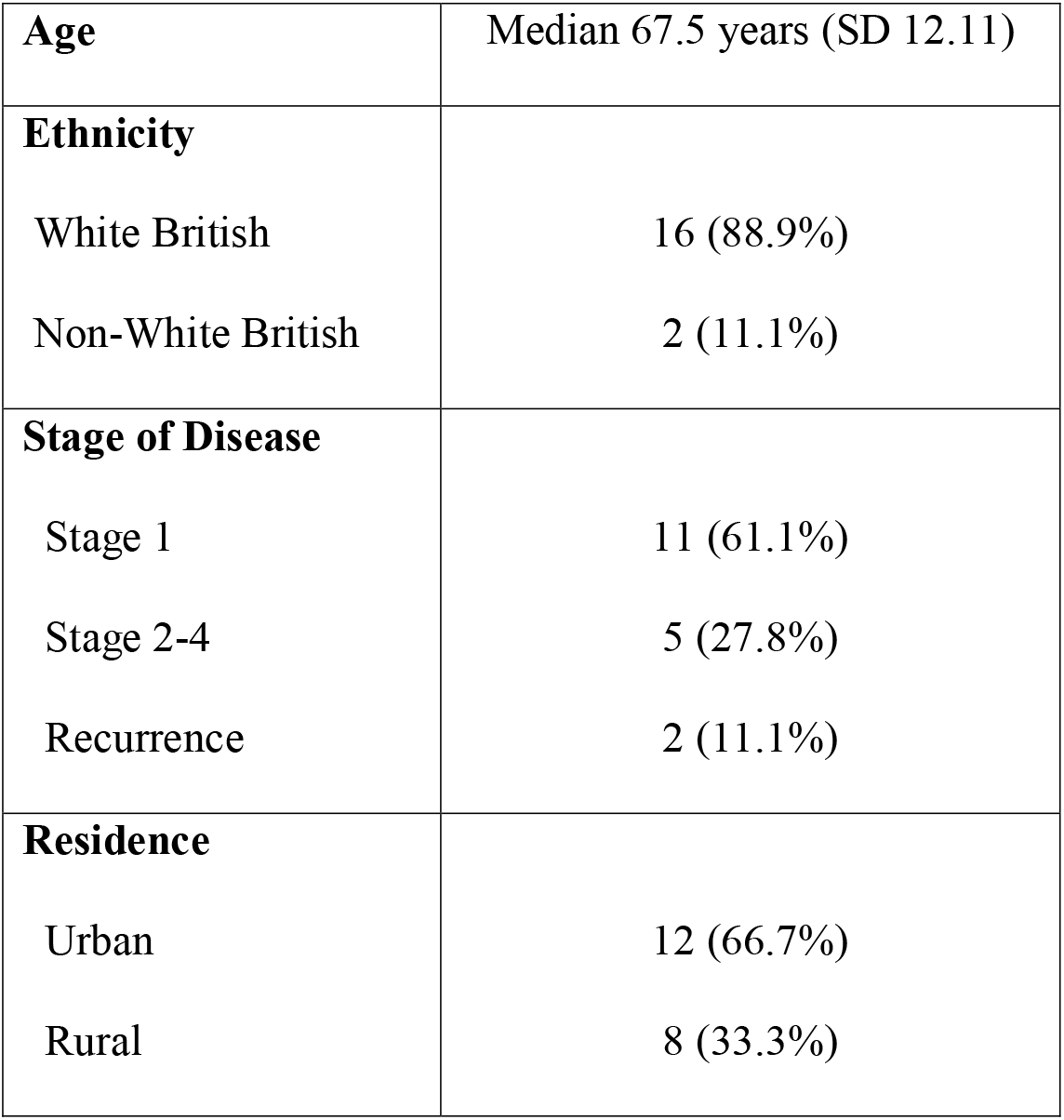
Participant characteristics, n=18.

|  |  |
| --- | --- |
| **Age** | Median 67.5 years (SD 12.11) |
| **Ethnicity** |  |
| White British | 16 (88.9%) |
| Non-White British | 2 (11.1%) |
| **Stage of Disease** |  |
| Stage 1 | 11 (61.1%) |
| Stage 2-4 | 5 (27.8%) |
| Recurrence | 2 (11.1%) |
| **Residence** |  |
| Urban | 12 (66.7%) |
| Rural | 8 (33.3%) |

### Thematic structure

Five main themes emerged from the data 1) Motivation for taking part in the study; 2) Experiences taking part in the study; 3) Patient perceived utility and acceptability of ctDNA in EC; 4) Follow-up experiences and preferences; and 5) PIFU and ctDNA as a potential follow-up tool. There were numerous subthemes identified within the main themes (Supplementary data).

### Motivations and experiences participating in the ctDNA monitoring study

Altruism was the leading motivation for study participation with a desire to help women in the future, even though they may not personally benefit from the results of the study. The participants shared their understanding of the purpose and potential role of the ctDNA blood test, the majority demonstrating sound understanding by discussing it as a new tool for cancer detection and how ctDNA blood testing could enable cancer recurrence to be detected earlier.

All participants regarded blood tests as having high patient acceptability, regardless of any possible discomfort. Even participants who had experienced minor discomfort as part of the blood testing process regarded that discomfort as acceptable and manageable, *‘I don’t mind it – I don’t mind, no, even if she can’t get the blood out [laughter], I just say ‘that’s alright, just try’…’ – 02*. Multiple participants made reference to difficulties in administering the blood test in an understanding manner, viewing it as a minor inconvenience and not problematic.

### Patient perceived utility and acceptability of ctDNA

Participants did express apprehension in relation to the ctDNA blood test, but this was only in relation to the lead-time for blood test results to become available. A clinical examination gave an immediate result and the apprehension expressed by participants may suggest that patients could feel concern or worry during the time between having blood taken for ctDNA testing and being given the results.

*Yes, that’s the thing, then you’ve got to wait for your blood test [result], and you’d think ‘oh I hope that’s alright’*.*’ – 02*.

Surprisingly participants raised very few concerns about false negative or positive results with the ctDNA blood test. This was summarised by one participant:

*‘If say it’s 80% plus accurate I think that’s reasonable but if you say it’s only 50% accurate, I think it could then cause problems and doubts about it*.*’ – 15*,

Participants also acknowledged that a clinical examination had its limitations.

The lead-time of a ctDNA test detecting EC recurrence potentially many months before symptoms developed was discussed by participants at length. Women acknowledged that a raised result may cause anxiety, and possibly even panic, at the thought of disease recurrence. Participant stated:

*‘You might feel you have a ticking time bomb’ - 16*.

Participants highlighted the importance of appropriate clinical support and timely further investigations. Several participants held the view that the earlier a recurrence was detected the sooner it could be treated with one female stating:

*‘Well, if it’s detected then hopefully something can be done about it*.*’-14*.

During the interview, a scenario was presented to participants where cancer recurrence was detected by ctDNA before a lesion was visible radiologically. Some women felt that they would place more reliance on the result of imaging rather than a blood test. This was outlined by one participant:

*‘I’d probably have mixed thoughts, because I’d put my faith in the CT’ – 10*.

Despite this, the overwhelming opinion expressed was that they would want to know if their ctDNA level rose, one participant stated:

*‘Well, I think I’d want to know I think, you know that, because obviously they’re going to check on you anyway aren’t they, so I think I would sooner know*.*’-17*.

The rationale for this viewpoint was mixed, some women reporting that it would help them be more aware of symptoms, being able to plan for life events and others made comparisons with prostate-specific antigen (PSA) monitoring for prostate cancer. This is summed up by one participant as follows:

*‘think about the men with the prostate cancer, they’ve all got numbers and they all sit around and talk about “Well my number’s higher than yours,” it’s like some sort of game they play. But they know it’s there and they know they’re being checked, so I don’t think it’s really that much different in the long run, yes*.*’-15*.

A key theme that emerged when discussing the potential role of ctDNA in detecting disease recurrence was ‘personal willingness to have a blood test to monitor for cancer recurrence, instead of a clinical (pelvic) examination’. Participants expressed an overwhelming preference for the blood test over a pelvic examination. One participant stated

*‘Oh yes. Oh yes! [laughter]… Rather have a blood test any day*.*’ – 01*, with further participants adding

*‘…it would be a lot easier wouldn’t it*.*’ – 04*.

Multiple participants laughed during this response, and this may signify agreement with an obvious answer. Many participants spoke about finding a pelvic examination difficult, pointing out that the examination was manageable but not psychologically acceptable. In the most part, participants expressed high levels of pragmatism and stoicism in relation to pelvic examinations, justifying any potential discomfort or embarrassment in order to monitor for and avoid cancer recurrence. One participant described the pelvic examination

*‘…of course you’re embarrassed, obviously, but you know it’s got to be done don’t you. You have to get on with it…’ – 09*.

A small number of participants described a severe reaction to the need to have a pelvic examination. One participant stated

‘*It still like puts the fear of God into me if someone said they wanted to examine me down there*..*’ – 18*.

One participant raised the issue of being examined by different doctors as compared to having a blood test taken by different people

*‘you have to brace yourself again because you will have an examination, quite possibly somebody you’ve never met before*..*’-16*.

### Follow-up experiences and preferences

Participants reported that regular HFU appointments had enabled them to ask questions and discuss any problems or concerns with their doctor, leading to a feeling of doctors being ‘*there for them’*, which provided a sense of reassurance. One participant described:

*‘…they benefit my health and frame of mind, because if I’m at all concerned, I can say that I’m concerned and I can be reassured’. – 07*.

However, participants also stated that the main downside to regular hospital follow-up was feelings of concern or worry when attending hospital appointments, linking to the second-level theme of ‘apprehension leading up to hospital appointments’. Participants were divided into feeling ‘apprehensive’ or ‘stoical’ about appointments. One female described how she felt about attending the hospital for follow up:

*‘…I think it’s just the thought of coming into hospital. As a patient… There is always that little niggle at the back of your mind that there might be something else, I think… it’s an unknown entity isn’t it…’ – 06*

Continuity of care was an issue for several participants and the wish to see the same doctor at each follow-up appointment:

*‘I do like to know that the person I’m talking to knows me, and knows what I’ve gone through and where I am and that with my treatment*…*’ – 01*.’

### PIFU and ctDNA as a potential follow-up tool

Participants gave mixed responses regarding PIFU as an alternative follow-up model to HFU, with some women expressing a preference for seeing a doctor face-to-face. Most participants however, expressed confidence in their ability to phone for advice if using a scheme such as PIFU. A few stated that they may ‘put off’ calling, out of a desire to ‘not bother’ the team,:

*‘I don’t like to bother people, I’m one of those people that, ‘well I’ll leave it, I’ll see how it goes…’ – 02*.

Other potential positives of PIFU were discussed, including the reassurance of having direct access to their clinical team, as well as practical benefits in particular of saving time and travel to appointments. One participant stated:

*‘it’s time as well, I’m back at work part time… you’ve probably got to change your shift or whatever, you know so in that respect it would be easier*.*’ – 04*.

Participants also stated that being trained on the red-flag symptoms to look for as part of PIFU could make them more confident, physically self-aware, and more able to self-check. One participant stated:

*‘That would be good, because as long as I was well-informed on what to look for…cos I mean I know they can’t carry on seeing me forever*.*’ – 07*.

Participants gave mixed responses about their personal willingness to be transferred to a PIFU scheme, however, all participants shared that they would be willing to be followed-up using a combination of PIFU and ctDNA monitoring. Having an ongoing medical test, such as a ctDNA blood test, would alleviate concerns of cancer recurrence and make PIFU acceptable as a follow-up tool. One participant gave a detailed explanation:

*No that’s good… I’d be quite happy… if I thought I was fine, and I was having a blood test every six months, that confirmed I was fine, that’s super you know, give me the little plastic*

*envelope and I’ll take it into my GP… and get a phone call back or a letter back… ‘thank you very much for your blood test, it was fine, come and see us in six months’, have another one in six months, that’s fine…’ – 01*

And a further participant stated:

*‘To be able to just go in and have a blood test, and them say ‘yes you’re alright, no you’re not’, that would be good. ‘Less time wasted, confidence that you have got the backup and confidence that hopefully the blood test is accurate and could discover something earlier than a hospital visit, definitely*.*’-15*.

## DISCUSSION

In this study we were able to explore the experiences and opinions from a diverse population in order to gain a wide breadth of experiences of women in active follow-up for EC. The results have identified new insights that can be used to inform the design of future trials incorporating ctDNA monitoring into clinical practice. The accuracy of ctDNA to detect EC recurrence in the pilot data from our study was 100% [12] and other studies support our results, including a lead time of over 6 months and high negative predictive value, 100% overall survival in gynaecological cancer cases with undetectable ctDNA [16].

One of the main findings of this study was the high level of patient acceptability of ctDNA monitoring, with an overwhelming preference by participants for a blood test rather than a pelvic examination to monitor for recurrence. The level of distress that was reported to be associated with a pelvic examination was considerable in some participants and sheds new light on a routine aspect of gynaecological practice in this population. The use of a diagnostic examination that is associated with such high levels of anxiety in some women, can result in patients entering a passive status, leading to feelings of powerlessness and helplessness [17].

Feeling unable to control or influence their health status may result in decreased levels of self-efficacy, and an external locus of control [18, 19], thereby reducing a patient’s ability to manage symptoms and perform maintenance behaviours [18]. A pelvic examination was identified as the *‘most personal’* of examinations, and although the majority of women accepted that it was a necessary aspect of their follow-up examination, some did raise the issue of continuity in the clinician-patient relationship in improving its tolerability, as compared to anyone performing venepuncture. A proportion of women may find pelvic examination completely unacceptable, either due to physical discomfort, a previous distressing experience or sexual violence, and by declining what is regarded as a mandatory component of their follow-up may experience even greater levels of fear of recurrence and guilt, should their cancer recur, due to a possible delay in diagnosis. In contrast, a ctDNA blood test was described as a psychologically acceptable alternative by participants, with any minor discomfort manageable and not problematic. A more acceptable and less invasive diagnostic examination would give a better experience, thereby empowering patients and prevent feelings of powerlessness and helplessness.

Current guidelines [20, 21] do not mandate regular imaging as part of clinical follow-up and therefore the ability of a clinician to detect distant metastases, particularly in the presence of obesity, is limited. PORTEC-3 reported that the site of the majority of first recurrences occurring in high-risk EC were distant, 176 cases (95.1%), compared to only 2 isolated vaginal recurrences (1.1%), and isolated pelvic recurrences, 7 cases (3.8%) [4]. The imperative for early identification of local EC recurrence lies in the ability to salvage such cases with radiotherapy, thereby improving overall survival, and supports the need for clinical examination. However, the majority of women who develop a local EC recurrence will present clinically, typically with vaginal bleeding [22]. Direct access to the specialist team through a PIFU scheme [8] may expedite clinical review, rather than patients taking a more passive role in their care and waiting until their next HFU appointment to report symptoms, as happens with a proportion of patients, possibly leading to a delay in diagnosis [22].

The women recruited to this interview study were already under HFU and participating in a cohort study determining the sensitivity of ctDNA to detect recurrence, but were blinded to their results, and therefore had previously received information on ctDNA monitoring as part of the ECctDNA study recruitment process. Despite women acknowledging that early detection of recurrence would be associated with anxiety, great importance was placed on being informed of the result, rather than being blinded and only the clinician being aware. The concept of remote biomarker monitoring for cancer is well established in many solid tumours, for example prostate and thyroid cancer, and has been shown to be well tolerated by patients [23]. Significant cost savings to both the patient and the healthcare economy have been identified with PIFU [10], as compared to HFU, and extension of such schemes could be used to finance the addition of ctDNA monitoring for EC follow-up.

Fear of cancer recurrence (FCR) can have a very real psychological impact on cancer survivors [24] and has been estimated to affect between 22% to 99% of all cancer survivors [25]. In a systematic review [25], a number of factors were found to be strongly associated with FCR including cancer cues, including new symptoms, pain and follow-up appointments. Some participants in our study expressed feeling anxiety when attending for follow-up appointments, in keeping with the review findings. The theme of ‘reassurance’ was stated as the most important benefit of HFU, reducing fears of recurrence, and helping patients to feel ‘looked after’, mirroring findings that regular hospital check-ups can provide patients with psychological reassurance and relief [26]. HFU was nevertheless associated with negative feelings in some women, linked to concerns over pelvic examination or the fear of disease recurrence, as well as practical issues of time and transport costs. The need for additional reassurance with PIFU was echoed in the views of our participants, however the addition of ctDNA monitoring was felt to be highly acceptable since it fulfilled the wish for disease monitoring, avoided pelvic examination but also gave them a greater sense of self-efficacy with regard to their health.

## Conclusions

This study provides in-depth insights into patients’ views on the acceptability of ctDNA for monitoring of EC, with women stating an overwhelming preference for a blood test instead of a pelvic examination. Participant insights also revealed that PIFU in conjunction with ctDNA, would be an acceptable approach to EC follow-up, supporting the proposal of a randomised trial comparing patient and healthcare benefits as compared to traditional HFU schemes.

## Data Availability

All data will be made available upon publication

## Author contribution

EM, HM and DG devised the study. AC and DG participated in conducting the ECctDNA study. AR conducted and analysed the interviews with support from EM and HM. AR and EM wrote the manuscript and all authors approved the final version.

